# Drivers of Decision-Making for Adult Tracheostomy for Prolonged Mechanical Ventilation: A Qualitative Study

**DOI:** 10.1101/2024.01.20.24301492

**Authors:** Anuj B Mehta, Steven Lockhart, Allison V Lange, Daniel D Matlock, Ivor S Douglas, Megan A Morris

**Author notes:** **Corresponding Author**: Anuj B Mehta, MD. 777 Bannock St. E320. Denver, CO, 80204.

## Abstract

**Background:** Decision-making about tracheostomy and prolonged mechanical ventilation (PMV) is emotionally complex. Expectations of surrogate decision-makers and physicians rarely align. Little is known about what surrogates need to make goal-concordant decisions. We sought to identify drivers of tracheostomy and PMV decision-making.

**Methods:** Using Grounded Theory, we performed a qualitative study with semi-structured interviews with surrogates of patients receiving mechanical ventilation (MV) being considered for tracheostomy and physicians routinely caring for patients receiving MV. Recruitment was stopped when thematic saturation was reached. Separate codebooks were created for surrogate and physician interviews. Themes and factors affecting decision-making were identified and a theoretical model tracheostomy decision-making was developed.

**Results:** 43 participants (23 surrogates and 20 physicians) completed interviews. A theoretical model of themes and factors driving decision-making emerged for the data. Hope, Lack of Knowledge & Data, and Uncertainty emerged as the three main themes all which were interconnected with one another and, at times, opposed each other. Patient Wishes, Past Activity/Medical History, Short and Long-Term Outcomes, and Meaningful Recovery were key factors upon which surrogates and physicians based decision-making. The themes were the lens through which the factors were viewed and decision-making existed as a balance between surrogate emotions and understanding and physician recommendations.

**Conclusions:** Tracheostomy and prolonged MV decision-making is complex. Hope and Uncertainty were conceptual themes that often battled with one another. Lack of Knowledge & Data plagued both surrogates and physicians. Multiple tangible factors were identified that affected surrogate decision-making and physician recommendations.

**Implications:** Understanding this complex decision-making process has the potential to improve the information provided to surrogates and, potentially, increase the goal concordant care and alignment of surrogate and physician expectations.

**Highlights:**

- Decision-making for tracheostomy and prolonged mechanical ventilation is a complex interactive process between surrogate decision-makers and providers.
- Using a Grounded Theory framework, a theoretical model emerged from the data with core themes of Hope, Uncertainty, and Lack of Knowledge & Data that was shared by both providers and surrogates.
- The core themes were the lenses through which the key decision-making factors of Patient Wishes, Past Activity/Medical History, Short and Long-Term Outcomes, and Meaningful Recovery were viewed.
- The theoretical model provides a roadmap to design a shared decision-making intervention to improve tracheostomy and prolonged mechanical ventilation decision-making.

## Introduction

In the United States, nearly 100,000 adults undergo tracheostomy annually, mostly to enable prolonged respiratory support.[1, 2] While a tracheostomy can facilitate life-prolonging interventions like mechanical ventilation (MV), it is also associated with significant morbidity and mortality. The majority of patients with a tracheostomy require prolonged hospitalization and intensive rehabilitation.[2, 3] Older adults with a tracheostomy often have a median survival of three to six months with high rates of readmission, frequent complications, and prolonged hospital stays.[3–5] The struggle between the potential for longer survival versus the complications associated with prolonged life support can make tracheostomy-related decisions emotionally complex for surrogate decision-makers.

A simple view of decision-making would be that physicians provide data to surrogates and surrogates make the final decision. However, previous studies suggest that decision-making for tracheostomy is far more complex.[6, 7] Surrogates want physician input and view (or want) the process to be collaborative. However, existing data suggests that there is significant dissatisfaction with decision-making in critical care settings. Previous studies have shown that surrogate and physician expectations about tracheostomy outcomes rarely align.[6] Significant disagreement exists in the literature with some reports indicating that most patients would find being attached to a machine “worse than death”, while other studies suggest patients are satisfied after a tracheostomy.[8, 9] Patients and families feel uninformed and that their values were not considered.[7, 10–12] The net result is that patients and surrogates are deeply unsatisfied with the decision-making process.

Shared decision-making (SDM) is the collaborative process of patients, surrogates, and providers reaching an informed collective agreement on the treatment most consistent with a patient’s values and is recommended as a core part of care for critically ill patients.[13–17] A necessary first step in SDM is understanding the decisional needs of key stakeholders. However, large gaps exist in understanding surrogate value structure and decisional needs as well as physician’s mental frameworks for making recommendations. These gaps may contribute to previous failed attempts at improving decision-making and reducing decisional conflict.[18] We conducted a qualitative decisional needs assessment to build a model for tracheostomy decision-making by identifying factors influencing surrogates and physicians.

## Methods

### Please see the Online Supplemental Methods for full details

#### Study Design

We conducted a qualitative study of surrogate decision-makers of patients being considered for tracheostomy and critical care physicians routinely involved in tracheostomy decisions from 2018-2022. Standards from the Consolidated Criteria for Reporting Qualitative Research were followed (See **eTable 1)**.[19] Grounded Theory methodology was used throughout the study.[20, 21] Grounded Theory aims to develop an explanatory theory of processes by understanding conceptual categories of importance related to the primary research question and can help build a model of a specific phenomenon.[20, 21]

#### Participants

Surrogate decision-makers were recruited from two hospitals and critical care physicians were recruited from multiple academic, public, and private physician practices. Surrogates were eligible if they represented a patient being considered for tracheostomy (see **Online Supplement** for details), were ≥18 years, and were English-speaking. Up to three surrogates per patient were allowed to enroll.[22] Critical care physicians who routinely care for patients receiving MV and engage with surrogates about tracheostomy related decisions were recruited through invitational emails.

A convenience sampling approach was initially used, followed by theoretical sampling to increase the number of surrogates who decided not to pursue a tracheostomy and physicians in private practice. Participant recruitment was discontinued once thematic saturation, the point at which no new themes emerge with additional interviews, was reached.[23, 24] Multiple interruptions to recruitment occurred due to the COVID-19 pandemic.

#### Data Collection and Analysis

Separate surrogate and physician interview guides were developed, and semi-structured interviews were conducted in person or virtually by study members trained in qualitative interviews (**Online Supplement**). Interview transcripts were coded and categorized using a constant comparative method and inductive approach.[25] The first five transcripts in each group were independently double-coded to develop separate codebooks for surrogates and physicians. Thereafter, every fifth transcript was double-coded to ensure calibration. An open team-based coding process allowed conceptual themes to emerge from the data.

Atlas.ti v9 (Berlin, Germany) was used for data management. All participants provided informed consent for the study. The study was approved by the National Jewish Health Institutional Review Board (HS-3136) and the Colorado Multiple Institutional Review Board (20-3102).

## Results

### Participants

Forty-three participants (23 surrogates and 20 physicians) participated in interviews (**Table 1**). Surrogates had a mean age of 48.2 years (SD=15.4) and were predominantly either the spouse/partner (26.1%) or child (34.8%) of the patient. Physicians had a mean age of 45.1 years (SD=7.5) with a broad range of years in practice.

**Table 1:** Participant Characteristics.

|  | Surrogate (n=23) | Physician (n=20) |
| --- | --- | --- |
| Age (mean, SD) | 48.2 (15.4)<sup>A</sup> | 45.1 (7.5) |
| Female, n (%) | 16 (76.2)<sup>A</sup> | 4 (20) |
| Race, n (%) |  |  |
| White | 9 (39.1) | 15 (75.0) |
| Black | 9 (39.1) | 2 (10.0) |
| American Indian/Native American | 2 (8.7) | 0 (0) |
| Other/Unknown | 3 (13.1) | 3 (15.0) |
| Hispanic, n (%) | 4 (17.4) | 2 (10.0) |
| Relationship to Patient (%) |  | NA |
| Spouse/Significant Other | 6 (26.1) |  |
| Child | 8 (34.8) |  |
| Sibling | 3 (13.0) |  |
| Other/Unknown | 6 (23.1) |  |
| Where do you practice? (%) | NA |  |
| Academic Setting |  | 8 (40.0) |
| Private Practice Model |  | 4 (20.0) |
| Hybrid Model |  | 8 (40.0) |
| How long have you practiced critical care medicine? (%) | NA |  |
| <5 years |  | 4 (20.0) |
| 5-9 years |  | 5 (25.0) |
| 10-14 years |  | 7 (35.0) |
| >15 years |  | 4 (20.0) |

Abbreviations: SD – standard deviation. n – number.
<sup>A</sup>2 surrogates did not provide demographic information. No missing data was imputed.

### Theoretical Model

Based on the interviews, a theoretical model of tracheostomy-related decision-making emerged from the data (**Figure 1**). Three conceptual themes were identified: Hope, Uncertainty, and Lack of Knowledge & Data. Additionally, four tangible factors were identified as contributing to surrogate decision-making and physician recommendations about tracheostomy: Patient Wishes, Past Activity/Medical History, Short and Long-Term Recovery, and Meaningful Recovery. Meaningful recovery meant different things to different participants but as a concept, arose repeatedly in interviews. Decision-making existed as a complex balance between surrogates and physicians with the factors affecting their decisions/recommendations being viewed through the lens of the conceptual themes. Decision-making was a bidirectional process with provider comments, opinions, and prognosis weighing heavily on surrogate decisions. Surrogates did not view the provider’s role as simply offering information and the final decisions resting solely with the surrogate. An additional layer of complexity emerged in the way in which themes and factors were interrelated (e.g., the Lack of Knowledge & Data contributed to Uncertainty for nearly all participants). From the surrogate perspective, the interplay between the themes contributed to their underlying value structure and was key to guiding the overall decision-making process.

**Figure 1.**
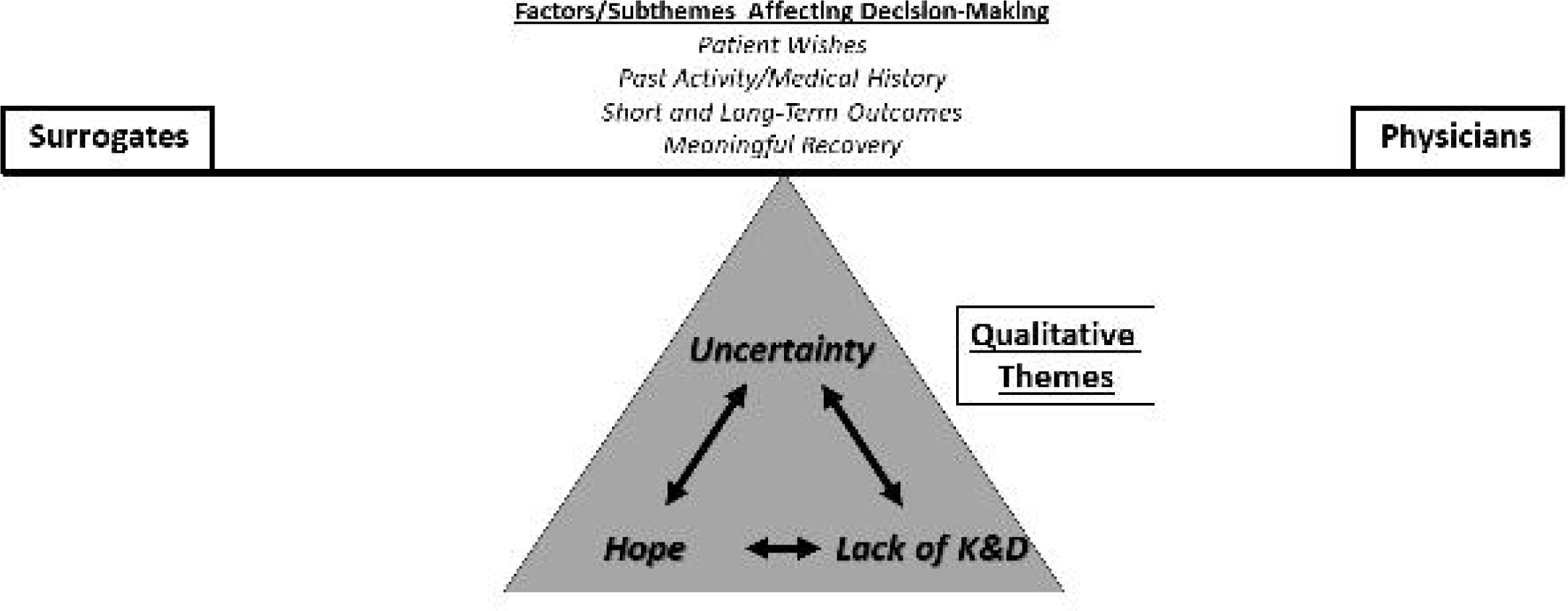
Tracheostomy Decision-Making Theoretical Model: Based on interviews with surrogates decision-makers and physicians, a theoretical model emerged from the data. Three conceptual themes where identified: Hope, Lack of Knowledge & Data (Lack of K&D), and Uncertainty. The bidirectional arrows between these themes indicates the significant interconnectedness of these themes. In addition to the conceptual themes, four tangible factors also emerged: Patient Wishes, Past Activity/Medical History, Short and Long-Term Outcomes, and Meaningful Recovery. The conceptual themes were the lens through which the factors were evaluated by both surrogates and physicians. The theoretical model depicts the conceptual qualitative themes as being the fulcrum of decision-making and that the process is a balance between surrogates and physicians influenced by the factors. Abbreviations – Lack of K&D – Lack of Knowledge & Data

### Hope

Hope was one of the most common concepts discussed by surrogates, anchoring most decision-making **(Table 2, eTable 2**) Hope was the lens through which many surrogates viewed past medical issues and influenced considerations about short- and long-term outcomes and meaningful recovery. When considering different possible outcomes, Hope sometimes superseded factual understanding of past medical issues (e.g., impact of chronic oxygen dependence from pulmonary fibrosis on the chances for ventilator liberation) or even the potential for recovery. Some surrogates indicated that despite knowing the high percentages of poor outcomes, they still held onto Hope for recovery. Some surrogates connected the feeling of Hope to their faith, “We just believe that prayer and everybody’s faith and hope is there. And it’s just a different way of looking at life I guess than a lot of the medical field looks at it, and we never knew that.” (*Surrogate ID1)*

**Table 2:**
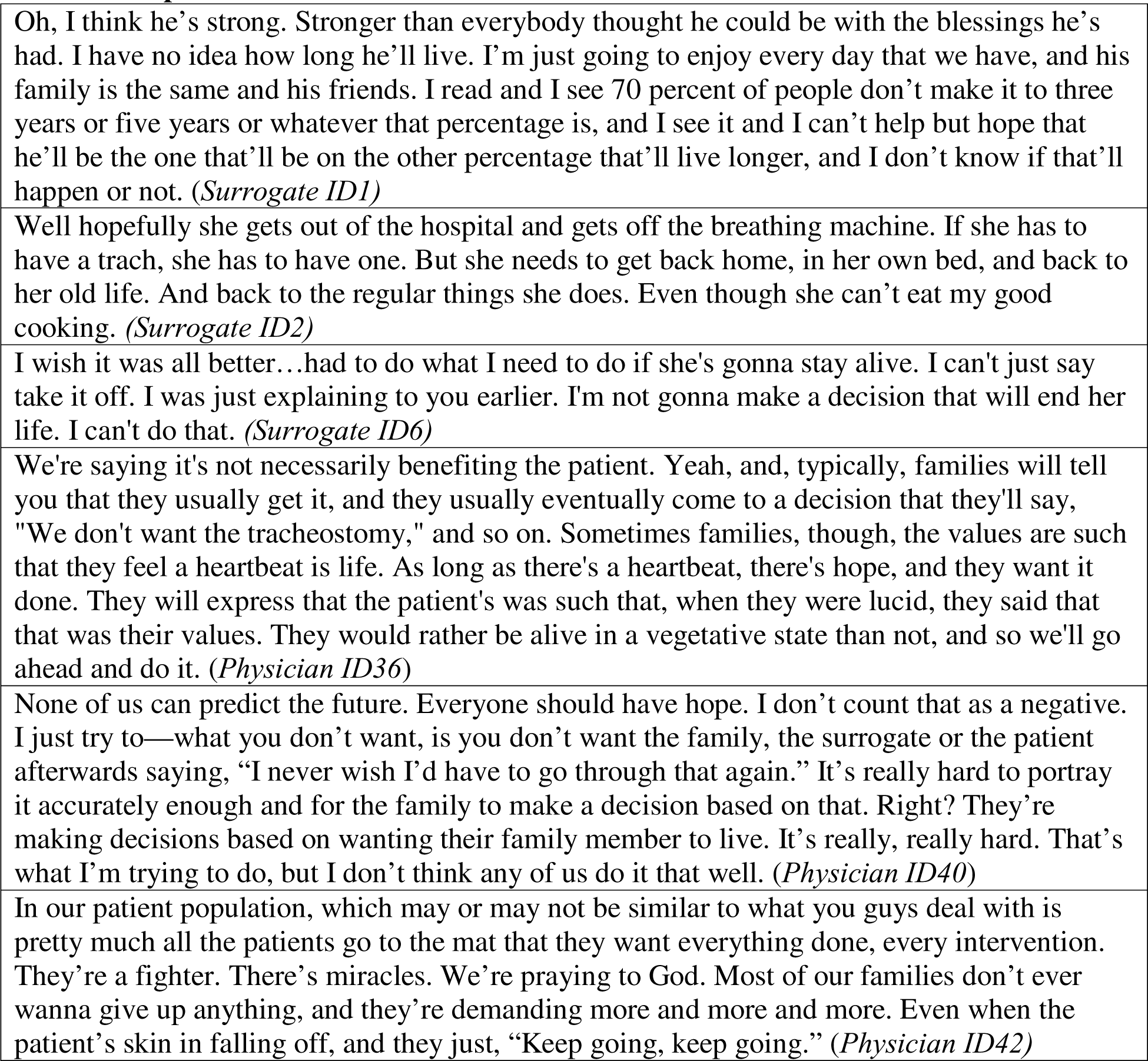
Hope.

|  |
| --- |
| Oh, I think he's strong. Stronger than everybody thought he could be with the blessings he's had. I have no idea how long he'll live. I'm just going to enjoy every day that we have, and his family is the same and his friends. I read and I see 70 percent of people don't make it to three years or five years or whatever that percentage is, and I see it and I can't help but hope that he'll be the one that'll be on the other percentage that'll live longer, and I don't know if that'll happen or not. (*Surrogate ID1*) |
| Well hopefully she gets out of the hospital and gets off the breathing machine. If she has to have a trach, she has to have one. But she needs to get back home, in her own bed, and back to her old life. And back to the regular things she does. Even though she can't eat my good cooking. (*Surrogate ID2*) |
| I wish it was all better...had to do what I need to do if she's gonna stay alive. I can't just say take it off. I was just explaining to you earlier. I'm not gonna make a decision that will end her life. I can't do that. (*Surrogate ID6*) |
| We're saying it's not necessarily benefiting the patient. Yeah, and, typically, families will tell you that they usually get it, and they usually eventually come to a decision that they'll say, "We don't want the tracheostomy," and so on. Sometimes families, though, the values are such that they feel a heartbeat is life. As long as there's a heartbeat, there's hope, and they want it done. They will express that the patient's was such that, when they were lucid, they said that that was their values. They would rather be alive in a vegetative state than not, and so we'll go ahead and do it. (*Physician ID36*) |
| None of us can predict the future. Everyone should have hope. I don't count that as a negative. I just try to—what you don't want, is you don't want the family, the surrogate or the patient afterwards saying, "I never wish I'd have to go through that again." It's really hard to portray it accurately enough and for the family to make a decision based on that. Right? They're making decisions based on wanting their family member to live. It's really, really hard. That's what I'm trying to do, but I don't think any of us do it that well. (*Physician ID40*) |
| In our patient population, which may or may not be similar to what you guys deal with is pretty much all the patients go to the mat that they want everything done, every intervention. They're a fighter. There's miracles. We're praying to God. Most of our families don't ever wanna give up anything, and they're demanding more and more and more. Even when the patient's skin is falling off, and they just, "Keep going, keep going." (*Physician ID42*) |

Physicians spoke about Hope from a different perspective. Many physicians described struggling with the cognitive disconnection between surrogates’ Hope for meaningful recovery and the clinical probability for recovery for patients with severe comorbid disease like cancer or acute illness like cardiac arrest: “I think family members who end up having tracheostomy put in patients who probably have no hope of getting better are, in a sense, kicking the can down the road, meaning they’re not—they’re hoping God intervenes and makes ‘em better.” (*Physician ID37).* When physicians felt that there was little Hope for meaningful recovery (e.g., older patients, patients with severe chronic comorbidities, or those with poor baseline functional status), they would often steer conversations away from tracheostomy.

### Lack of Knowledge & Data

For surrogates, the Lack of Knowledge often was related to the specific factors affecting decision-making identified in the theoretical model especially short- and long-term outcomes and meaningful recovery (**Table 3, eTable 3)**. Surrogates mentioned feeling uninformed about the likelihood of ventilator liberation, regaining the ability to eat or talk, regaining functional status, returning home, and overall survival. One surrogate was unclear about what the “quality of life” would be after a tracheostomy or any potential alternatives even after the discussion with the medical team: “Can they have a quality of life? Can they do things? Or are they just bound to a bed and that machine? Is there a portable machine that helps them breathe or is it – I don’t know. What options would there be? *(Surrogate ID3)*. Multiple surrogates indicated that the Lack of Knowledge about rehabilitation and long-term acute care facilities and associated costs made tracheostomy decisions much more difficult, “I think that I’m still a little fuzzy about the long-term. Because they had said that he was going to go to a long-term rehab facility. His insurance doesn’t cover that.” *(Surrogate ID18)*. Other surrogates indicated that even after discussing tracheostomies with the physicians, they were still unaware of potential alternative pathways. Some surrogates also struggled with Lack of Knowledge about a patient’s basic medical information either due to lack of health literacy or poor communication, “He may have had shortness of breath and maybe passed out and had been unconscious… You know what, I think they did say he had a cardiac arrest” (*Surrogate ID23*).

**Table 3:** Lack of Knowledge & Data.

|  |
| --- |
| Maybe more details about the rehabs and absolutely about the rehabs, what he'd do there, what the possibility – we were wanting to ask about if he can't walk, would he have to have a mobile chair, wheelchair or those little gadget chairs that move around, what can he do to still have a little bit of quality of life to get around. We wanted to ask if the dry weather was going to be better than humid weather, warm weather better than cold weather altitudes, but they – we never got a chance to sit down. And even when we were in the team meetings, we weren't ever asked do you have any questions ever. (*Surrogate ID1*) |
| Interviewer: Sure. What kind of education would want in terms of the different things that people can do with a trach, a tracheostomy?<br>Interviewee: Can they have a quality of life? Can they do things? Or are they just bound to a bed and that machine? Is there a portable machine that helps them breathe or is it – I don't know. What options would there be? (*Surrogate ID3*) |
| Educational stuff. What you can expect. . . .Going forward with it. The amount of sicknesses. Even if you don't go over every sickness that you can get because yeah, you can get everything from not having a trach too. But the chances of getting sick go up. The chances of having a respiratory infection go up. But really what you can expect is the biggest thing that I would really have wanted to know going forward with somebody with a trach. (*Surrogate ID7*) |
| But I think if the trach is seriously being considered, if there's a good prediction tool that could really say, "There's a better chance than not this is a permanent fixture to this person's body, and it may mean they're gonna be living in the nursing home forever," that can be helpful. (*Physician ID28*) |
| I think that would be good information to tell patients, here's where this patient fits in terms of expected outcomes and trying to come to that decision with the family. Is it really worth it for us to pursue the tracheostomy or not? I think if we had a good data that showed us this patient fits this phenotype and they have this percentage chance to come off the ventilator, I think that would be very beneficial. . . . I try not to do that, but if I had objective data that I could plug the specific patient's information into and say, "Hey, 50 percent of patients have this outcome," then I think that—or to stay on the ventilator for this long when they have this particular course to this particular disease pathology. I think that's good information for them to manage their expectations and say, "Hey, maybe this is worth it, maybe this isn't," so that just everyone's more informed. (*Physician ID39*) |

Additionally, surrogates described a Lack of Knowledge about the impact of past medical issues on recovery, with one surrogate not appreciating that chronic oxygen use was a major health issue prior to the acute illness, negatively impacted chances for recovery. Surrogates also expressed a Lack of Knowledge about a patient’s wishes, even those who had severe illness prior to coming to the hospital: “I think it’s difficult to make decisions for somebody. I know that I know her for a long time, but this is something we never actually talked about” *(Surrogate ID20)*. Several providers also highlighted that the Lack of Knowledge about possible outcomes and alternatives, even after family meetings, negatively impacted surrogates’ experience and led to significant decisional conflict: “But I think they also need to know what the alternative is. I think they a lot of times make a decision to proceed with tracheostomy based on fear of the alternative, as opposed to wanting the tracheostomy.” (*Physician ID28*).

For physicians, the corollary of Lack of Knowledge was Lack of Data, specifically around being able to accurately predict short and long-term outcomes and meaningful recovery. The majority of physicians highlighted the Lack of Data around time spent in rehabilitation facilities, ability to be liberated from the ventilator, and overall survival as negatively impacting their confidence in recommendations provided to surrogates. Most physicians also discussed the potential for meaningful recovery, often described as a return to previous physical function combined with the ability to interact with loved ones, as distinct from other outcomes. Meaningful recovery arose inductively as a key factor affecting physician recommendations, “I think someone who doesn’t have a bridge therapy to a good quality of life, so say someone who has a cardiac arrest and it has pretty obvious anoxic brain injury and there’s not likelihood for meaningful recovery, I try to steer the family away from a tracheostomy.” (*Physician ID39*)

The Lack of Knowledge & Data also led participants to state that more education and decision-support tools may improve the process, “Education stuff is really what I would want” (*Surrogate ID7*). Some physicians even indicated that decision-support tools might offer greater benefit to providers as it might standardize discussions around tracheostomy especially when there was a Lack of Data: “I think that would be very helpful. I think they [surrogates] want numbers that I feel like I can’t provide them. What is his chance of coming off the vent? I end up really pulling numbers out of just from general sense of my past experience, which not good. ‘Cause everyone’s so different. It’s just if we could get those numbers, it’d be awesome… But I think that would be great.” (*Physician ID27*).

### Uncertainty

Uncertainty manifested as an intangible feeling about the entire decision-making process described by both surrogates and physicians (**Table 4, eTable 4**). Uncertainty was intricately linked to Hope and Lack of Knowledge & Data. For many surrogates, decision-making was described as a battle between feelings of Hope and feelings of Uncertainty. Moreover, while Lack of Knowledge & Data dealt with concrete information and outcomes, it fueled the general sense of Uncertainty that surrogates and physicians described as contributing to decisional conflict. Patient wishes were reported to be important to both surrogates and physicians but views on these wishes were often influenced by Uncertainty even for patients with severe comorbid disease, “Because of the fact that he didn’t have any advance directive, we were winging it” *(Surrogate ID7)*. Surrogates with a clearer view of a patient’s past wishes reported being less burdened during the decision-making process. Physicians also expressed their own Uncertainty about what might happen after a tracheostomy when discussing matters with surrogates. Physicians’ Uncertainty often revolved around more general concepts about the potential for recovery, the temporary or permanent nature of the tracheostomy, etc., “it’s [recovery] uncertain. Right? So, I can’t actually tell ‘em, but I can say … I say, “I can’t guarantee anything in medicine with the exception of I can guarantee we’re honest. I’ll tell you if they got worse or better… I’m not gonna get too high or too low.” (*Physician ID33*). One of the most common ways in which Uncertainty manifested for physicians was when they felt that patients had a poor chance of meaningful recovery but because of Uncertainty about outcomes, patient wishes, or surrogate values, they reported still recommending tracheostomy to surrogates. In connection with a Lack of Data, physicians also described Uncertainty when presenting two sides of a potential outcome without much information to help surrogates see which was more likely (e.g. some patients coming off the ventilator quickly while others may need the tracheostomy and ventilator for a long time).

**Table 4:** Uncertainty.

|  |
| --- |
| So I've often wondered, . . . Because, of course, you don't want to give up on your family member at all, especially when you're the one making the decision. You don't want to make that decision. But it's just hard because you can't foresee the future. (*Surrogate ID5*) |
| I think it's difficult to make decisions for somebody. I know that I know her for a long time, but this is something we never actually talked about (*Surrogate ID20*) |
| Interviewer: Okay, has that been hard not knowing what your dad would've wanted if he got really sick?<br>Interviewee: Well, it's just hard bein' in that position of knowing or not knowing, but having to make that decision [laughter] for him as far as what to do health wise, and it's extremely uncomfortable. I believe that that was maybe the hardest part in this whole thing, not just because I didn't know what my dad would want, but also because I am a firm believer in Jesus Christ. I didn't want to play the role of God in any decision-making like that, though, as far as life and death. (*Surrogate ID22*) |
| Interviewee: I don't know at first . . . I guess I was just worried. I was worried if he would be able to breathe on his own, and then I was also worried as far as how long they would be allowed to leave him on there 'cause I wasn't really sure how that worked, if they could only leave him on there for a certain amount of days, and then take him off.<br>Interviewer: Gotcha, so that uncertainty?<br>Interviewee: Mm-hmm. (*Surrogate ID23*) |
| I guess it's usually tailored to the clinical situation, but there's always uncertainty, so I'll usually—the hope would be that this would be a short-term thing and I guess that's one other thing that I usually add on to your prior question which if this is reversible? You can take it out and the hole will heal up and go away, usually within days. . . . I think it's, hopefully, temporary, but it's always hard to know. May often translate into leaving the hospital to go to a long-term acute care hospital, so different physical settings. Sometimes that can be seen as at least an encouraging sign of recovery to make it out of the hospital, but also to know that it could be something that goes on for days, weeks or even months. In rare cases, it's clear that it's going to be indefinite, in which case I'll certainly try not to pull any punches here in these discussions and I'll let them know if I think it will be indefinite. (*Physician ID35*) |

Additional participant quotes describing can be found in **eTable 5-8**.

## Discussion

In a prospective qualitative study, we identified conceptual themes and key factors contributing to surrogate and physician decision-making for tracheostomy. Using Grounded Theory, a theoretical model of tracheostomy decision-making emerged (**Figure 1**).[20, 21] The themes were the lens through which the more tangible factors were viewed and processed. Decision-making represented a balance between surrogate needs and physicians’ ability to provide recommendations. Unlike classic conceptions of decision-making where providers give information to surrogates and surrogates are left to make decisions on their own, this study revealed that tracheostomy and PMV related decision-making is a highly interactive process between surrogates and providers.

Current decision-making approaches for tracheostomy, prolonged MV, and other critical care interventions are inadequate for surrogates, with poor alignment between surrogate and physician expectations.[6, 12] Xu et al reviewed family meetings and identified similar themes to our study including past wishes and long-term prognosis but surrogates and physicians were not directly interviewed by unbiased researchers.[26] Other studies with pediatric populations also found similar themes to this study (e.g., functional status, past medical history, prior wishes, etc.).[27, 28] Given the emotional complexities of tracheostomy decision-making, the lack of a theoretical model may contribute to the lack of efficacy of past decision-support tools for prolonged mechanical ventilation.[18] To our knowledge, this study is one of the first to use qualitative methodologies to build a theoretical model for adult tracheostomy decision-making.

In traditional Grounded Theory, themes emerge during the analysis process that can then be used to build a theoretical model of the phenomenon being studied.[20, 21] For tracheostomy decision-making, the themes reflected the surrogate’s value structure or the value structure of the patient as perceived by the surrogate. However, tracheostomy decision-making is emotionally complex and frequently encompasses larger goals of care and end-of-life discussions. It often involves weighing quality of life with quantity of life. As in other areas of health care, this balancing act can be influenced by concrete factors such as the patient’s past wishes, past medical history, and the potential for meaningful recovery. Therefore, the theoretical model that emerged from the data included both conceptual themes and factors that exist as a balance between surrogate views and physician recommendations.

A key finding was the interaction between the themes. Surrogates logically recognized that pre-existing conditions like cancer or chronic oxygen use might impact the chances for recovery but in many cases, Hope overrode data driven decision-making. Lack of Knowledge & Data was common finding for both surrogates and physicians. The Lack of Knowledge & Data referred to concrete factors such as a lack of information about rehabilitation facilities, data on the chances for ventilator liberation, etc. Sometimes, it appeared that the Lack of Knowledge & Data gave surrogates more Hope since no data was viewed as better than bad data. However, Lack of Knowledge & Data also fueled Uncertainty for surrogates and physicians.

While Uncertainty was one of three themes to emerge, it was also central to all other themes. In 2011, Han et al described a new taxonomy for Uncertainty given its key importance in health care.[29] Han et al described multiple sub-types of Uncertainty in health care including prognostic Uncertainty, Uncertainty related to structures and processes of care, and existential Uncertainty. These subtypes of Uncertainty encompass many of the themes and factors identified in this study. Prognostic Uncertainty aligns with Lack of Knowledge & Data. Uncertainty related to structures and processes of care was akin to surrogate concerns about rehabilitation options. Existential Uncertainty also mirrored the classic intangible form of Uncertainty identified as a core theme in this study. This deeper understanding of Uncertainty may highlight some of the core struggles for surrogates and physicians.

SDM is recommended by multiple medical societies and expert groups as a core component of complex decision-making including for decisions like tracheostomy in the ICU.[10, 14, 16, 30, 31] A core part of SDM is defining a patient or surrogates underlying value structure to best present potential outcomes. Providing more data to surrogates does not always improve decision-making, especially for surrogates who lack numeric literacy. However, when the possible risks, benefits, and outcomes presented to surrogates align with their personal value structure, behavioral theory would suggest that the additional data may reduce internal conflict.[32] In fact, most surrogates and some physicians stated that the lack of information and education on alternatives, rehabilitation options, and the likelihood of different outcomes negatively impacts the decision-making process. Moreover, a lack of understanding of the surrogate decision-making process also contributes to wide variation in physician approaches to discussing tracheostomy and goals of care. This study may provide a more concrete framework from which to approach future SDM interventions.[18]

This study has multiple limitations. Despite transitioning to a theoretical sampling approach towards the end of the study, it remained difficult to enroll surrogates who had chosen not to pursue tracheostomy. While many expressed interest, not pursuing tracheostomy was often accompanied by the patient dying and surrogates struggled to commit to interviews while grieving. As such, there is a selection bias. Additionally, the COVID-19 pandemic caused a nearly two-year interruption in recruitment as research was paused at an institutional level and because tracheostomy practices evolved during the first year of the pandemic. While the study was multi-center, different themes may influence decision-making in other regions and institutions related to different populations and different hospital structures. Finally, surrogates and physicians were interviewed separately and not as part of a dyad. Therefore, it was not possible to determine what was actually said during tracheostomy conversations, only surrogate recollections of what was said. However, surrogate experience and final decisions are always based in perceptions of decision-making conversations making the themes identified in this study highly relevant to improving the decision-making process.

The theoretical model for tracheostomy decision-making demonstrates a complex interplay between the qualitative themes of Hope, Lack of Knowledge & Data and Uncertainty. These themes were the lens through which the more concrete drivers of decision-making (patient wishes, past activity/medical history, short- and long-term outcomes, and meaningful recovery) were viewed. The model also reveals that surrogates view the provider role as being much larger than just offering information or fact sharing. While some have argued that the physician’s role is to only provide information and that final decisions rest on the patient or surrogate, our findings indicate that decision-making a is bidirectional process.

The gaps identified by the theoretical model represent areas where additional outcomes data may aid the decision-making process but also highlights the importance of clarifying patient/surrogate value structure through which to present such data. This study highlights the complex interplay between surrogates and physicians and how a collaborative approach to decision-making is needed. The current theoretical model can serve as the foundation for SDM tools designed to improve goal-concordant care.

## Site

The work was a joint effort between Denver Health and Hospital Authority, the University of Colorado School of Medicine, and National Jewish Health

## Data Availability

All data produced in the present study are available upon reasonable request to the authors and pursuant to local and federal regulations.

## Acknowledgements

ABM is supported by NIH K23HL141704 (Primary funding source). ABM had full access to all the data in the study and takes responsibility for the integrity of the data and the accuracy of the data analysis. Contents are the authors’ sole responsibility and do not necessarily represent official NIH views. No authors had any conflicts of interest related to this study. The funders did not have any role in design and conduct of the study; collection, management, analysis, and interpretation of the data; preparation, review, or approval of the manuscript; or the decision to submit the manuscript for publication.

## Author Contributions

ABM, ISD, and MAM conceived the study. ABM and SL were responsible for data collection and analysis. ABM, SL, and MAM were responsible for data interpretation. ABM drafted the article. ABM, SL, AVL, DDM, ISD, and MAM provided critical revisions and meaningful input for the final draft. ABM had full access to all the data in the study and takes responsibility for the integrity of the data and the accuracy of the data analysis. ABM conducted all aspects of data analysis. All authors approved the final draft of the manuscript.

