## Supplement for "Drivers of Decision-Making for Adult Tracheostomy for Prolonged Mechanical Ventilation: A Qualitative Study"

**Online Supplement**

**Table of Contents**

1. Online Supplemental Methods
2. eTable 1: Consolidated Criteria for Reporting Qualitative Research (COREQ)
3. Final Surrogate Interview Guide
4. Final Physician Interview Guide
5. eTable 2: Additional Quotes for Hope
6. eTable 3: Additional Quotes for Lack of Knowledge & Data
7. eTable 4: Additional Quotes for Uncertainty
8. eTable 5: Additional Quotes for Patient Wishes
9. eTable 6: Additional Quotes for Past Activity/Medical History
10. eTable 7: Additional Quotes for Short and Long-Term Outcomes
11. eTable 8: Additional Quotes for Meaningful Recovery

**Online Supplemental Methods**

*Study Design*: We conducted a prospective qualitative study using semi-structured interviews recruiting surrogate decision-makers of patients being considered for tracheostomy and physicians routinely involved in tracheostomy decisions from 2018-2022. The participants were surrogates of patients being considered for tracheostomy and critical care physicians who routinely care for patients with respiratory failure. Surrogates and physicians were recruited separately. Participants were recruited from 2018-2022. There were multiple interruptions to participant recruitment due to funding gaps as well as the COVID-19 pandemic. At an institutional level, research was halted for prolonged periods of time in 2020 and 2021. Additionally, tracheostomy practices changed dramatically during the early phase of the pandemic and decision-making did not reflect usual practice.

*Surrogate Recruitment*: Surrogates were recruited from two institutions in an urban metro area. The first institution had was an academic community hospital with a single combined medical and surgical ICU. While labeled as a “community hospital”, this institution also served as the regional referral hospital for complex respiratory failure for an entire health system. Surrogates were also recruited from an academic urban safety-net hospital with a dedicated medical ICU. Surrogates were recruited from two ICUs in an urban area. One ICU was a mixed medical and surgical unit in an academic community hospital that served as the referral center for a hospital system. The other ICU was a dedicated medical ICU located in an academic safety-net hospital that also had a large referral encashment.

Surrogate decision-makers were recruited in a two-step process. Surrogates were first identified if they represented a patient who was considered “eligible for tracheostomy” although the patients themselves were not study participants. A patient was considered “eligible” if they (1) were ≥ 18 years of age and (2) receiving MV for ≥ 7 days or if tracheostomy was already being considered by the medical team based on chart review. Patients were excluded if they (1) did not have a surrogate decision-maker, (2) were pregnant, (3) were incarcerated, (4) had chronic respiratory failure due to a progressive neuromuscular disease, (5) were being considered for tracheostomy due to trauma or airway obstruction, (6) had a high suspicion of death within 48 hours not related to transitions to comfort measures, or (7) were expected to be extubated within 72 hours based on the primary medical team’s clinical assessment. The study team screened patients on a daily basis and if a patient was thought to be potentially in need of a tracheostomy, a study member confirmed eligibility with the primary medical team and obtained permission to contact the surrogate decision-maker. Surrogates were approached for participation if they were (1) ≥18 years of age, (2) identified by the primary medical team as the surrogate medical decision-maker for a patient receiving MV, and (3) spoke English. Surrogates were excluded if (1) they did not speak English, (2) could not provide informed consent, or (3) were incarcerated.

*Physician Recruitment*: The target physician population was critical care physicians who routinely care for patients receiving MV and have discussions with surrogates about tracheostomy and PMV. It was not possible to recruit surrogate-physician dyads as each physician recruited cared for more than one patient being considered for tracheostomy. Additionally, the goal of the study was to explore the decision-making process. Decision-making decisions typically occur in a family meeting settings with the primary critical care team, not necessarily the proceduralist who will do the tracheostomy. As such, proceduralists who only do the final consent but are not involved in the earlier decision-making process were not recruited. Specific inclusion criteria included (1) age ≥18 years, (2) worked routinely in a critical care unit caring for patients with respiratory failure, and (3) had been in practice for at least one year. Physicians were excluded if they had not been in practice for at least one year.

Physician recruitment took two forms. Invitation emails were sent to divisional email lists from institutions where study members had academic, research, or clinical affiliations or relationships. Additionally, a snowball method whereby participants were able to refer other colleagues to the study team was used.

*Qualitative Methods and Analysis*: Standards from the Consolidated Criteria for Reporting Qualitative Research (COREQ) were followed and details can be found in the **eTable 1**.^1^ A Grounded Theory framework was used for the study design and analysis.^2,3^ Grounded Theory is a qualitative methodology that posits that meaning is derived by understanding structural processes. Grounded Theory develops an explanatory theory of processes by understanding conceptual categories of importance related to the primary research question and can help build a mental or theoretical model of a specific phenomenon.^2,3^ Utilizing this framework to build a mental model of decision-making, separate interview guides were designed for surrogates and physicians (**Online Supplement**). Individual interviews or group interviews for patients with more than one surrogate decision-maker were conducted in person or virtually by study members trained in qualitative interviews (AM and SL).^4^ A convenience sampling approach was initially used. However, the study eventually transitioned to a theoretical sampling approach to attempt to increase the number of surrogates who decided not to pursue a tracheostomy as well as the number of physicians in private practice. Participant recruitment was terminated once thematic saturation had been reached. Thematic saturation is the point at which no new themes or concepts emerge with additional interviews.^5,6^

The transcripts were coded and categorized also using a Grounded Theory analytic framework with an inductive approach.^2^ The first five surrogate transcripts and first five physician transcripts were dual coded (AM and SL) to develop a separate codebooks for surrogates and physicians. After the codebook was finalized, every fifth surrogate and fifth physician transcript was dual coded to ensure calibration. An open team-based coding process was used to allow themes to emerge from the data. In addition to real-time coding as interviews were conducted, a constant comparison approach was also used to compare data from completed transcripts to newly completed interviews to inform the categorization of themes and concepts.^7^ During the analytic process, the study team met regularly to discuss emergent new codes and themes, and assess preliminary and final results.

Atlas.ti v9 (Berlin, Germany) was used for data management. All participants provided informed consent for the study. The study was approved by the National Jewish Health Institutional Review Board (HS-3136) and the Colorado Multiple Institutional Review Board (20-3102).

**eTable 1: Consolidated Criteria for Reporting Qualitative Research (COREQ)**

| **Domain 1: Research Team and Reflexivity** | | |
| --- | --- | --- |
| **1** | Interviewer/Facilitator | ABM, SL |
| **2** | Credentials | ABM – MD, MS  SL – MPH  AVL - MD  ISD – MD  DM – MD, MPH  MAM – PhD, MPH |
| **3** | Occupation | ABM – critical care physician, researcher  SL – senior qualitative analyst  AVL – critical care fellow, palliative care physician, critical reviewer  ISD - critical care physician, researcher, site lead  DM – geriatric physician, expert in Shared Decision-Making  MAM – qualitative methodologist and researcher |
| **4** | Gender | Male – ABM, SL, ISD, DM  Female – AVL, MAM |
| **5** | Experience and Training | ABM – intermediate qualitative experience, coursework, workshops, extensive critical care experience  SL – extensive qualitative experience,  AVL – intermediate qualitative experience, coursework, workshops, moderate critical care experience  ISD – minimal qualitative experience, extensive critical care experience  DM – extensive qualitative and shared decision-making experience  MAM – extensive qualitative experience, methodologist qualitative and mixed methods research core |
| **6** | Relationship established | The study was introduced to all participants prior to enrollment. ABM had clinical responsibilities at all sites. |
| **7** | Participant knowledge of the interviewer | The goal of understanding decisional needs around tracheostomy was made clear to all participants. No deception was used. All interviewers presented their credentials and role in the study to each potential participant. |
| **8** | Interviewer characteristics | Participants were aware of the interviewer credentials and qualitative experience. They were aware of the PIs research goals and biases inherent in the PI being a critical care physician. They were also made aware of the funding for the project. |
| **Domain 2: Study Design** | | |
| **9** | Methodological orientation and theory | Grounded theory |
| **10** | Sampling | Convenience sampling initially but then a transition to theoretical sampling to increase the number of surrogates who refused tracheostomy and physicians in private practice. |
| **11** | Method of approach | Email, face-to-face, flyers, and phone calls |
| **12** | Sample size | 45 |
| **13** | Non-participation | 11 people expressed interest but were unavailable for interviews. |
| **14** | Setting of data collection | In-person interview in conference room settings initially that transitioned to entirely virtual interviews via web-based conferencing system or phone |
| **15** | Present of non-participants | Only researchers and participants were present |
| **16** | Description of sample | See Table 1 |
| **17** | Interview Guide | See eTable 2 and 3 for the final version of the interview guide. The interview guides were developed with input from key stakeholders. |
| **18** | Repeat interviews | No |
| **19** | Audio/visual recordings | Audio recordings were made for all interviews. |
| **20** | Field notes | Field notes were taken for all interviews and focus groups. |
| **21** | Duration | The stated goal for focus groups and interviews was less than 60 minutes for surrogates and less than 30 minutes for physicians. A few interviews exceeded these time limits. |
| **22** | Data saturation | Data saturation was discussed among the research team and ensured through data analysis |
| **23** | Transcripts returned | No |
| **Design 3 Analysis and Findings** | | |
| **24** | Number of data coders | 3 – ABM and SL |
| **25** | Description of coding | An open, inductive, team-based process was used to develop separate codebooks for both surrogate and physician interviews. The first five surrogate and first five physician transcripts were independently double coded and then reconciled until consensus was reached and there was strong code agreement to develop separate codebooks. Thereafter, every fifth transcript for surrogates and physicians were double coded (20% of all transcripts) to ensure calibration. Any discrepancies were discussed and resolved by consensus. Themes from surrogate interviews and physician interviews were identified. |
| **26** | Derivation of themes | Themes were derived from the data |
| **27** | Software | Atlas v9 (Berlin, Germany) |
| **28** | Participant checking | Themes were informally reviewed with a random sample of participants (member-checking) and informally discussed with several ICU practitioners not involved in the research (triangulation). |
| **29** | Quotations | Yes |
| **30** | Data and findings consistent | Consistency was present between raw data and extracted themes/findings. |
| **31** | Clarity of major themes | See Results, Table 2-4, eTable 4-6. |
| **32** | Clarity of minor themes | See Results, Table 2-4, eTable 4-6. |

ABM – Anuj B Mehta MD. SL – Steven Lockhart MPH. AVL – Allison V Lange MD. ISD - Ivor S Douglas MD. DM - Daniel Matlock MD MPH. MAM - Megan A Morris SLP PhD MPH.

**Final Surrogate Interview Guide**

1. Patient’s Past
   a. Tell me about the your loved one before they came into the hospital.
      i. What types of activities does he/she enjoy?
      ii. *Where does he/she live?*
      iii. *Does he/she work, go to school, or are they retired?*
   b. Briefly tell me about some of their chronic major medical issues prior to this hospitalization.
   c. Have you ever talked to them about what they would want if they were very sick?
      i. *Have they ever had an advanced directive or living will? If so, do you know what it said?*
2. Medical Course
   a. Can you tell me a little about the medical issues that brought your loved one into the hospital?
   b. Can you tell me about the communication you have received from the medical team?
      i. *Who has been giving you the most information?*
      ii. *Tell me about things you liked about the communication process.*
      iii. *Tell me about things you would change about the way in which the medical team communicated with you.*

3. Tracheostomy Discussion
   a. This study is about tracheostomy decision-making but not everyone has had a full discussion about a tracheostomy with the medical team. Can you tell me a little about what you know about a tracheostomy and how anything related to tracheostomy has been communicated to you?
      i. *Who first discussed it with you?*
      ii. *When did they discuss it with you?*
      iii. If it has been raised an option for [patient], can you tell me why the medical team thought it was an option to consider?
      iv. How did they describe it to you?
   b. Can you tell me about your feelings about [patient] being attached to a breathing machine for a long time?
   c. Can you tell me a little bit about some of the options for [patient] if they cannot come off the breathing machine (i.e. cannot breathe on their own) soon? What options has your medical team discussed with you?
      i. *Do you think you understood all of the options?*
   d. What types of questions come to mind when you are talking with the medical team about the breathing machine and the possible need for a tracheostomy and long term support from the breathing machine? Were they answered?
   e. If you have talked to the medical team about a tracheostomy, do you feel that you have been appropriately educated about what it is, why it might be needed, and what to expect long-term?
   f. *Did you feel prepared to make the decision after talking with them?*
   g. *Did you feel the medical team had a preference?*
4. Tracheostomy Decision
   a. Have you made any decisions about tracheostomy and long term mechanical ventilation? If so, can you tell me about your decision?
   b. If you have not made any decisions, tell me about what things you would consider when making a decision about tracheostomy. What additional information might you want?
   c. Which members of the medical team should be involved in discussions and decisions about a tracheostomy (e.g. physician, RN, RT) ?
   d. What considerations or factors went into or would go into making the decision?
      i. *What the medical team recommended?*
      ii. *What you think your loved one would want?*
      iii. *What might happen with a tracheostomy?*
           1. *Being attached to a breathing machine, being in a nursing facility for a long time, eating, talking ,etc.*
   e. What types of information or discussions could make a tracheostomy decision easier? (e.g. educational information, information about long-term care facilities, predictions about short and long-term outcomes, individualized decision-aid)?
5. Tell me about how you see the near future for your loved one and your family?
   a. If you did not or do not choose the tracheostomy, what do you expect in the next few days (what alternative would you choose)?
   b. If [patient] does/did have a tracheostomy:
      i. Tell me about your expectations and goals for them.
      ii. Tell me about what you think about them coming off the breathing machine?
      iii. What did your medical team tell you to expect?
6. Has COVID-19 affected the way you think about being on a ventilator or life support for a long time? If so, how?
7. What recommendations would you give someone else making the decision about a tracheostomy for their loved one?

**Final Physician Interview Guide**

Physician Experience

1. Can you tell me a little about where do you practice?

Physician Practice

2. Can you describe to me the types of patients for whom you raise the possibility of a tracheostomy?
3. When do you tend to start thinking about a tracheostomy for a patient and what influences that thought process?
4. Are there specific types of patients for whom you tend not to discuss tracheostomy or for whom you recommend against a tracheostomy? Can you tell me about your thought process when deciding not to recommend a tracheostomy?
   a. Can you tell me about a recent patient who was on the ventilator for a “long” period of time where you either did not discuss or recommended against a tracheostomy?

Decision-Making and Communication

5. How do you approach families and surrogates for tracheostomy decision-making?
6. What do you tell them about a tracheostomy?
   a. *What do you tell them about the procedure itself?*
   b. Can you describe to me what you tell them about the possibility of prolonged life support?
7. How do you tell them what to expect in the short term and in the long term?
   c. Do you use any decision aids or prediction tools?
8. Can you describe to me how you think surrogates approach the decision-making process?
   d. *What do you think are the most important things that they want to know?*
9. What types of information do you think surrogates need to make tracheostomy related decisions?
10. Can you tell me a little about how you think surrogates perceive the decision-making process?
11. What do you think surrogates expect when they make a tracheostomy related decision?
    e. *Do they all assume that their loved one will get better or do they have a generally negative outlook?*
12. Do you think a decision aid or tailored prediction tools would help families?
    a. What would make a decision-aid easier for you to use?
13. How has COVID-19 affected the way you think about tracheostomy and prolonged mechanical ventilation?
    a. Has COVID-19 changed the way you make recommendations to families?

*Case Information*

14. *Tell me about a recent case in which you recommended a tracheostomy.*
15. *What issues arose with the surrogates during the decision-making process?*
16. *Tell me about what do you think could improve the decision-making process?*

**eTable 2: Additional Quotes for Hope**

I hope that he can—I don't know if he'll be able to go home in the same manner. I think he'll probably have to have some sort of—somebody, some home care. Somebody that stops by and checks on him and stuff like that. Whether he's gonna be able to drive and—I don't know. We'll just have to see how things go. *(Surrogate ID7)*

I’m scared that she’s not gonna have quality of life, but everything she has is something that you can control. Even down to the lymphoma, she’s young. It can be cured. The encephalitis, it can be cured [laughs]. The PRES, the main thing in it is that it’s reversible. We just don’t know, with all of them together, what it means. There’s hope because of all that, but there is unknown too. *(Surrogate ID12)*

Interviewer: I gotcha. I got it. Yeah. It sounds like as long as he’s not in pain, you want to give him the best shot he has, which may mean being on a breathing machine.

Interviewee: Who’s to say that he has to be on that machine all the time? It’s just for the moment right now. We live in the moment. We live for day by day, and that’s what we’re doing right now. We’re takin’ it day by day. *(Surrogate ID23)*

Well, I guess I'll always discuss it with the family. Patients that I would not recommend it to are the patients that have, before they were intubated, had a very poor quality of life. Like for instance a patient with end-stage dementia who then suffers a cardiac arrest, and there's really no reasonable hope that the patient's gonna regain any kind of quality of life. In that sort of instance, I would offer it, but I wouldn't recommend a tracheostomy. (*Physician ID27)*

…Quite often personal emotions get involved, though, and also kind of what they believe. Not all patients kinda believe the health care providers, and they don't always want to hear negative prognosis, and want to hold onto hopes for miracles and whatnot. So I think a lot of it is kind of personal versus what they would think that their family member would want. (*Physician ID26)*

Yeah. I don't do pediatrics, but so, yeah, not under 18 but, yeah, between—yeah, but between 18 and 30 is kind of the range, yeah, where we see the parents—typically parents and sometimes a spouse will really be, basically, making the decision to prolong the life over quality of life because there's a glimmer of hope. (*Physician ID36)*

**eTable 3: Additional Quotes for Lack of Knowledge & Data**

Interviewer: Did they talk to you about not doing the tracheostomy and taking the breathing tube out at that time, the last time?

Interviewee: No. *(Surrogate ID2)*

Interviewer: When you finally talked to the medical team about the tracheostomy, did they happen to tell you what to expect long term after a tracheostomy?

Interviewee: No *(Surrogate ID14)*

A few years ago, he was going through a bout of cancer. I don’t know a lot about what kind of cancer, or anything. I know that he went through treatments. I’m not sure if he’s in remission. We have a hard time with him being forward about what’s really going on with him. He has a hard time talking about it, doesn’t want to put people in a situation where they have to take care of him, or feel sorry for him. (*Surrogate ID18*)

…I think they [surrogates] want numbers that I feel like I can't provide them. What is his chance of coming off the vent? I end up really pulling numbers out of just from general sense of my past experience, which not good. 'Cause everyone's so different. It's just if we could get those numbers, it'd be awesome. I just wonder how applicable they would be to each individual case. But I think that would be great, yeah, to have. (*Physician ID27*)

Interviewer: Do you offer any specific predictions to family members about chances of coming off the machine, about being alive, you know, in a year, about needing to go back to the hospital and ongoing complications?

Interviewee: Oh, I don’t tend to. I just think it’s really tough to predict, you know? I think that it all depends on, you know, what rehab place they choose, you know, what rehab doctor they get, you know, what are, you know, the specific patient’s kind of ability to, you know, survive a prolonged stay so I’d say, you know, it’s pretty hard to predict that (*Physician ID30*)

…I don’t think there’s clear guidance in the literature about when you should do it and I think it’s more of a personalized decision…(*Physician ID33*)

I think if there's some uniformity that's based on evidence, that therefore would be reliable and you'd feel safe about giving these odds or these average measures of changes in recovery time or whatever it is. It's hard 'cause it's such a hugely diverse number of patients for different reasons that end up requiring tracheostomy. Anyway, I think it can only helpful…(*Physician ID35*)

To me, I don't know how important the actual decision about a tracheostomy is for a family-centered decision aid, but a family-centered decision aid about prolonged mechanical ventilation—what it is, what it entails, what are the outcomes associated with it, what would that life look like going forward—yeah, that would be useful. (*Physician ID43*)

**eTable 4: Additional Quotes for Uncertainty**

Interviewer: And before he went into the hospital, did you and he ever have conversations about if he were to get really sick potentially from this lung disease, but also from other things, about what he would want?

Interviewee: We might have talked about it, but I’ll tell you honestly, the amount of times we went to the clinic and this PCP’s, they presented nothing about what this could be to us. So it was a shock when – on [Date]. We had no inkling. . . So no, we didn’t discuss that before he went in the hospital (*Surrogate ID1*)

I thought about that [if loved one couldn’t come off breathing machine], but I have no idea. I guess she’d have to stay in the hospital and be in assisted living somewhere. I’ve been thinking about it. I don’t want to think about it, but it’s something I’ve thought about. *(Surrogate ID2)*

Interviewer: have you ever talked to him—did you guys ever discuss what they would want if they were very sick?

Interviewee: No. That's been one of the pains about this whole process is that he—this is just a conjecture, really, on my part. Bringing up the conversation was admitting I'm getting old, and I need to talk about it. He wasn't willing to converse too much about it. He was in the hospital about a year ago with a similar—it was more lung issues than it was heart issues as it was this time. Yeah. He just really didn't wanna discuss it, so unfortunately, no. *(Surrogate ID7)*

Well, if he be attached to a breathin’ machine for a long time, in the first place—this is what I talked with one of the doctors about—if they feel like he is not getting any better—‘cause I know you just can’t live on no machine—and I told them if they sees where he is not gettin’ any better and can’t survive without the machine, just take the machine off of him. Just let him just ease on and die on his own. I told them, don’t—somethin’ go to happen to him and don’t use no machine on him to try to save him or whatever. Just let him go on out. ‘Cause most times when they use that—left the machine on a person, they—the person already dead anyway. *(Surrogate ID24)*

So, yeah, well, if they have a poor prognosis for non-neurologic reasons, like if they had metastatic refractory, y'know, some sort of pancreatic cancer, something that was associated with a very poor prognosis. Or if they had a severe interstitial lung disease and I didn't think that there was really a chance that they would come off the ventilator at any point. Those are two other instances where I'd recommend against it, but still offer it. (*Physician ID27*)

Unfortunately it's [Tracheostomy] not a solution to the respiratory failure, and that can be sort of unpredictable in its resolution. 'Cause a lot of people will ask, "Well, how long are they gonna need this?" And that to me is kind of a moving target, or is a moving target in most patients. (*Physician ID28*)

I would say when I'm thinking of opening a tracheostomy there are people who the tracheostomy will help them get off the ventilator faster—maybe they might need less sedation and that sort of thing—or there are people who just need a long time to get off the ventilator. (*Physician ID43*)

**eTable 5: Additional Quotes about Patient Wishes**

… I know that the decision will be mine. I know that [patient] would not want to continue life in the state that he's in. I would have to make that decision. *(Surrogate ID3)*

…I wanted to make sure we were all on the same page of what we understood Dad wanted. And we have been. So that really helps, too. And his close friends that have been up here are in complete agreement… *(Surrogate ID5)*

I do know that my mom would want me to do everything that I could to save her. She doesn’t wanna be on life support. If the machine is keeping her alive? No. She doesn’t want that. She is a full code. If something happens, we are to try to keep her going, but not for months at a time. Like I said, if it’s life support and she’s got no brain activity *(Surrogate ID8)*

I always encourage them to try to follow not what they want to say, but what the patient would want. I think that kind of takes the onus off of them for making the decision to go to say comfort measures. Because there's nobody who wants to have that guilt of deciding to withdraw all care on a loved one, but if they think about if the patient would want that, quite often they realize that it would not be what the patient would want. (*Physician ID26*)

I kinda address what the trach can do to help but really until I understand what the patient’s wishes would be, what they would want in terms of long-term care, short term care, the trach takes a little bit of a backseat to that. (*Physician ID28*)

Families have a lot of emotional ties caught up in their tracheostomy decisions because sometimes they're associated with potential end-of-life decisions, right? If you don't go for the tracheostomy, you might be going for a terminal extubation …or what will likely be a terminal extubation. And because of that, I try to focus the decision on what is, you know, your loved one's goals of care? What would they want to be able to do and not do so that we start to bring the – keep the conversation focused around the patient, so that's one of the approaches I take. (*Physician ID34*)

It’s [having conversations with surrogates who do not know what patient wanted] really hard. There is a very rare patient, unfortunately, that we have really good advance care planning or saying, like, “Yep, if I get sick, I would like a trach.” That’s never really on those advance directives…It’s usually mechanical ventilation. The decision is like, “Do I be intubated or not?” Rare people have say, like, “I would like intubation or mechanical ventilation or support for 14 days. If I don’t turn around, I would like to focus on my comfort.” A lotta that has the decision of like, “Do I be intubated or not?” If you say, “Okay, yes, I do wanna be intubated,” it doesn’t provide a lotta guidance for families and surrogates to say where’s the limit to that. Yeah. That does get to be the challenge with a tracheostomy in particular ’cause we’re relying on families and surrogates and other decision makers to say yes or no, but it’s hard to have patient input on those decisions. (*Physician ID38*)

A lot of the times, you’re in the middle where the family—the person didn’t say. The family’s not sure. Again, the conversation really needs to be about the goals of the patient, and. if they don’t know, you just have conversations with the family to say, “Well, tell me about the person. Did they ever have a friend or another loved one who was in this situation, and what do you remember them saying about if that happened to me, what would I want?”

That can give you some clues about what they would want, yes or no. (*Physician ID42*)

**eTable 6: Additional Quotes about Past Activity/Medical History**

Interviewer: What are things she enjoyed?

Interviewee: She likes to go shopping, spending money. Likes to go to the movies. . . She did like bingo. Bingo was a big – it was very big for her. . . she loved being around everybody. She doesn’t like sitting at home by herself.

Interviewer: I understand she’s in a wheelchair, but can you describe to me about her limitations and taking care of herself before she came to the hospital?

Interviewee: Well pretty much what she can do is brush her teeth and put her makeup on. Now before she’s been having problems with that. She basically just moves pretty much one arm now. Other than that, she doesn’t do very much. She does get up and put one arm on the computer. We have a computer set up so she can – voice activated, so she can do it if she wants to. . . Her biggest peeve was trying to get – trying to keep CNAs to take care of her. That’s been a big problem since I don’t know . . . She needs the help. She’s always needed the help. Before then, I used to do it all. I can’t do it anymore either *(Surrogate ID2)*

Interviewer: Have you thought at all about what the possibilities would be if he couldn't come off the breathing machine?

Interviewee: To be honest with you, every day for the last six months, I fear that when I come home I'm going to find my husband dead. So, yeah, it's been a struggle. I think he's just given up on everything. I don't think he has the coping skills to deal with any of it. So, he's just drowning his sorrows. I feel like I'm helping to kill him because I know if he doesn't get the alcohol, that he's going to have another seizure. So, because he hasn't been able to get it on his own, I've been getting it for him. I'm feeling a huge amount of guilt about that. *(Surrogate ID3)*

So, yeah, well, if they have a poor prognosis for non-neurologic reasons, like if they had metastatic refractory, y'know, some sort of pancreatic cancer, something that was associated with a very poor prognosis. Or if they had a severe interstitial lung disease and I didn't think that there was really a chance that they would come off the ventilator at any point. Those are two other instances where I'd recommend against it, but still offer it. *(Physician ID27)*

In other words, for example if it's a 95 year-old with advanced COPD who I don't expect will ever – have an opportunity to get off the ventilator, I may focus my discussion on the fact that tracheostomy is a potential option. It may not be a great one in this setting. As opposed to the 35 year-old who had respiratory failure from influenza and a case of ARDS, and I expect things will gradually improve over time, though it may take a while, and that would – Ideally putting in a tracheostomy would make sense for long-term care of this patient while they recover *(Physician ID28)*

It’s not something I wouldn’t offer. I think if you have those patients who are old, has medical comorbidities or weak or deconditioned or have cancer or other things that are placing them at really high risk for complications for long-term mechanical ventilation, still we have that conversation with families. . . We rely a lot on families to have that conversation for the patient. That’s always a challenge because it’s like, “Does Grandma really wanna go to an LTAC? She really liked Bingo and liked being at home and drivin’ her car. She may have to move out of her house and liquidate all of her resources in order to afford or go to the LTAC or all these things.” Sometimes we don’t get to have a lot of patient input on those decisions (*Physician 38)*

If you have somebody who is metastatic cancer in five places, they’re on dialysis, they’re not awake or interactive, and they’re just never gonna get better, strokes on both sides, just a train-wreck patient, and they’re on a ventilator, again, it gets back to talking to the family about this person’s prognosis for life, irregardless of the trach, is not good. *(Physician ID42)*

**eTable 7: Additional Quotes about Short and Long-Term Outcomes**

Interviewer: And at any point, have you considered what would happen if she couldn’t come off the breathing machine, if her lungs weren’t strong enough?

Interviewee: I thought about that, but I have no idea. I guess she’d have to stay in the hospital and be in assisted living somewhere. I’ve been thinking about it. I don’t want to think about it, but it’s something I’ve thought about. *(Surrogate ID2)*

Comfortable wise, I think the tracheotomy is better for her and then she’s not gonna have all that stuff on her face. Yesterday, she did communicate with us by mouthing things. She told me she wanted to go home. Things like that. Now, she can mouth those things and we can actually see more of what she’s trying to say. *(Surrogate ID8)*

That the tube down her throat could—higher risk of infection and that after so many days, it becomes a much higher risk, and that the tracheostomy is temporary if needed. It could heal up on its own if it’s a short period of time. It could be sewn up. It could be used—you could take the breathing tube out. You could put the air in the oxygen type if needed. *(Surrogate ID9)*

Being on a machine or in a nursing home is just something that is gonna not sit very well with him. (*Surrogate* ID12)

Well, if he be attached to a breathin’ machine for a long time, in the first place—this is what I talked with one of the doctors about—if they feel like he is not getting any better—‘cause I know you just can’t live on no machine—and I told them if they sees where he is not gettin’ any better and can’t survive without the machine, just take the machine off of him. Just let him just ease on and die on his own. I told them, don’t—somethin’ go to happen to him and don’t use no machine on him to try to save him or whatever. Just let him go on out. ‘Cause most times when they use that—left the machine on a person, they—the person already dead anyway. *(Surrogate ID24)*

I have sort of my standard sort of tracheostomy things. I'll even say, "I know it sounds barbaric. It's a hole in the neck. But actually, patients will often find it more comfortable. (*Physician ID29).*

I’m not suggesting that people think this way, but they become less of like a sack of meat sitting on a bed and more of like a person where you actually look at their face. Saying like, “Oh my God. Look. That’s the way he looks like.” So, I’m actually not opposed to tracheostomy and all, but it’s typically when somebody looks like they’re making – it’s just gonna be gradual, slow improvement and they failed an extubation attempt once. *(Physician ID33*)

We do a couple things to try to remove their feeling of guilt if they're talking about prolonging life or if they're talking about . . . switching to comfort only. Typically, they will—we ask them to really consider what the patient would want. We know you love them, and you would want this or that. Of course, you would want them to live forever, but what would they want if I told them today that they're gonna be in the nursing home for the rest of their life, cared for other—by others for 24 hours, 7 days a week, that kind of thing? (*Physician ID36*)

I think they are universally surprised when they say they can’t talk and can’t eat. They don’t think that that’s—they think, “Oh, it moves down here. Now, my mouth works. I can do those things.” (*Physician ID41)*

I think that the subtleties of long-term tracheostomy outcomes are probably not well versed even in the critical care community…It’s a little bit different now in the post-COVID world, but for many, a trach was a means to the end of their initial ICU stay, that it’s like, “Okay, well, once they get trached and pegged” or whatever else, “they can go to an LTAC and see where it goes.” (*Physician ID45*)

**eTable 8: Additional Quotes about Meaningful Recovery**

Well, if I strongly disagree with their decision to proceed with a tracheostomy in a patient that I think has a poor chance of coming off the vent, then I try to just present them with the facts. Like, "Your loved one will be in a – most likely scenario is he or she will be in a chronic vent facility for an indefinite period of time, with a poor quality of life." *(Physician ID27)*

I really draw on personal experience from my work in the LTAC. . . So I'll explain what my observations have been in patients who have been sent to me while I'm covering the LTAC. What that looks like, who tends to recover, and who doesn't. *(Physician ID28)*

What brought them joy, because sometimes that really helps illustrate. Like say they were kinda of miserable at baseline and you've identified that this illness, they're never gonna fully recover from. Then, say that they're miserable at home. Are they gonna be more or less miserable in a nursing home, say? Forget about trach. I mean that would just be sort of a general, what's the least acceptable minimum—the minimally acceptable recovery for them? They would be willing to work through weeks or months of rehab to get to. *(Physician ID36)*

I'm pretty blunt in my discussions with the family. If I don't think it's gonna be a path to them getting better, meaning are we just essentially preparing them to be warehoused to live out their days in a long-term care facility or something like that, I don't necessarily discourage it, but I encourage the families to think about are we really benefiting your loved one or are we just delaying the inevitable, and that delay, essentially, affords this person no quality of life and no joy. *(Physician ID37)*

I think when you are able to paint the picture of exactly what the day-to-day would look like in a timeline and the fact that they’re gonna have to go to a nursing facility at some point. Even if they are able to come off ventilator and still have a trach in, they’re still likely gonna be very deconditioned. Would they want to spend the next several weeks to months in a nursing facility and still, even when they get discharged from that nursing facility—if they get discharged—there’s still gonna be a lot of long-term consequences with weakness. Their life will never be the same. *(Physician ID39)*
